# Covid19 infection spread in Greece: Ensemble forecasting models with statistically calibrated parameters and stochastic noise

**DOI:** 10.1101/2020.06.18.20132977

**Authors:** Georgios D. Politis, Leontios Hadjileontiadis

## Abstract

Following the outbreak of the novel coronavirus SARS-Cov2 in Europe and the subsequent failure of national healthcare systems to sufficiently respond to the fast spread of the pandemic, extensive statistical analysis and accurate forecasting of the epidemic in local communities is of primary importance in order to better organize the social and healthcare interventions and determine the epidemiological characteristics of the disease. For this purpose, a novel combination of Monte Carlo simulations, wavelet analysis and least squares optimization is applied to a known basis of SEIR compartmental models, resulting in the development of a novel class of stochastic epidemiological models with promising short and medium-range forecasting performance. The models are calibrated with the epidemiological data of Greece, while data from Switzerland and Germany are used as a supplementary background. The developed models are capable of estimating parameters of primary importance such as the reproduction number and the real magnitude of the infection in Greece. A clear demonstration of how the social distancing interventions managed to promptly restrict the epidemic growth in the country is included. The stochastic models are also able to generate robust 30-day and 60-day forecast scenarios in terms of new cases, deaths, active cases and recoveries.

## 1. Introduction

SARS Cov2 is a novel Coronavirus leading to infection of the respiratory system, namely Covid19, with a wide range of symptoms, including severe pneumonia and in some cases death, mostly in the elderlies and the individuals with prior health issues. On January 30^th^, 2020, WHO declared coronavirus outbreak a global pandemic. The following days, the virus started spreading across Europe and USA, resulting in a high number of fatalities during a very short period of time. By May 1^st^, 2020, more than 220,000 worldwide have passed away, while more than 750,000 have recovered from the infection.

In Greece, the government took early precautions so as to prevent a potentially fast epidemic spread along the country. Social distancing measures were applied since the first week of March and an official lockdown was declared on March 22. It is widely accepted that this policy resulted in a substantial reduction of the infection magnitude and the expected fatalities, while the local health system was able to effectively respond to the emergency. On April 27, the Greek state announced that the lockdown will be withdrawn on May 4^th^, but the social distancing measures will be maintained in a looser manner. Opening of senior and junior high schools was declared for May 11^th^ and 18^th^, respectively. By May 1^st^, Greece has confirmed 2530 cases of the new coronavirus and 139 fatalities.

In this study, statistically inferred compartmental models were developed in order to assess and forecast the evolution of the infection in Greece as well as the peak of the epidemic and the potential ending time of the first surge of the disease. 2 different sets of models were developed. The first model was developed in order to forecast the evolution of the first surge of the epidemic in Greece in terms of confirmed daily and accumulated cases and fatalities. The model produces a deterministic scenario alongside confidence intervals. The second model was developed in order to estimate the real magnitude of the epidemic in several stages of the social distancing measures as well as to produce probabilistic forecasts for the following month. Both models utilize the same pre-processing methodology, yet they are diversified in the next stages of this analysis, depending on the purpose each model serves. Thus, the first model is merely deterministic in nature, while the second is mostly stochastic and it ends up to a fully applicable ensemble forecasting model.

### 1.1. Relevant work

Numerous epidemiological studies have demonstrated the need to analyze the dynamics of infectious diseases, both in the context of steady state and transient characteristics. Most of them incorporate thorough timeseries analysis alongside spectral representation. Cazelles et al.,[1], used wavelet analysis to assess strongly non-stationary epidemiological data of various infectious diseases such as measles and cholera. The main aim of the study was to recognize long-term periodic patterns and identify spatial synchronies between different countries. In another study, Cazelles et al., [2], successfully reconstructed incidence data from flu by incorporating time varying parameters in an MCMC stochastic model. Lloyd, [3], revealed complex dynamical patterns in childhood viral diseases via the inclusion of realistic distributions of infectious period in a multistage SIR model. Krylova and Earn, [4], also used realistic distributions of the characteristic periods of infectious disease in order to reveal predicted transitions in patterns of measles dynamics.

Champredon et al., [5], mathematically demonstrated the equivalence of the renewal equation with the Erlang-distributed SEIR epidemic models. Wang et. al., [6], used a phase adjusted SEIR model in order to estimate the new Covid19 cases in China during the various stages of the social interventions. Particularly, they modelled the temporal variation of the reproduction number and then they implemented it to a simple SEIR model to infer the new cases. Van der Driesche, [7], provides simple methods to calculate the basic reproduction number of various infection diseases with or without control strategies included. Chae et.al., [8], used ARIMA models and deep neural networks for short-term forecasting of the spread of 3 infectious diseases, namely Chicken Pox, Scarlet Fever and Malaria. More particularly, they used social media data, as well as temperature and humidity as input variables to the NN models to predict the new incidences of each disease in South Korea.

From the review of the related work, it is evident that non stationarity testing and wavelet analysis have mainly been used for a long-term analysis of viral outbreaks, in an effort to capture long-term periodicity and seasonality. On the other hand, simple compartmental modelling and gamma distribution have mainly been used for short-term prediction. Short-term statistical forecasting has also been achieved through artificial intelligence. However, neural network forecasting requires big data. This study investigates the application of timeseries and wavelet analysis in a short-term prediction process in the first stages of a viral disease outbreak, when limited data is available and forecasting becomes an urgent matter for the healthcare providers.

## 2. Aim

The aim of the study is bi-directional. The primary aim is to provide insights into the realistic distributions of the epidemic in Greece as well as the characteristic times of the novel Coronavirus infection through an optimal combination of timeseries analysis and compartmental modelling. The secondary aim is to forecast the evolution of the outbreak in Greece with respect to the variations in social distancing measures. As it will be evident throughout the study, the developed models managed to forecast both the magnitude and the most critical stages of the first surge of the infection in the country.

## 3. Methodology

### 3.1. Definitions

The basic reproduction number is the number of secondary cases generated by a single case in a fully susceptible population. The incremental or effective reproduction number is the temporal variation of the basic reproduction number during the course of the epidemic spread and it can be estimated either with or without social interventions. The transmission ratio is a positive number in the (0,1) open interval, indicating the probability of secondary cases from a single case. It is proportional to the incremental reproduction number. The incubation period is the time duration between the exposure of an individual to the virus and the onset of symptoms. The recovery period is the time period between the onset of symptoms and the recovery/immunity. The infectious period is the time interval during which an individual can transmit the virus through the respiratory system. By definition, it is equal to the recovery period. However, under several circumstances, an individual can infect others during the pre-symptomatic stage. Also, interventions like quarantine and/or hospitalization after the onset of symptoms significantly reduce the infectious period due to lack of contact with other individuals. Both the incubation and the recovery period as well as the basic reproduction number are characteristic of the virus under study.

The CFR or case fatality rate is the proportion of the confirmed cases that pass away from the infectious disease under study. Usually, it differs from the actual fatality rate, due to the fact that not all individuals are tested and not all carriers of the infection are confirmed by testing.

The methodology is presented in the following steps:

### 3.2. Statistical pre-processing and basic parameters estimation

The spread of a respiratory infection in a population is a dynamic process characterized by substantial stochasticity. Subsequently, it becomes apparent that the timeseries of timestep cases and deaths of a viral disease result from a stochastic process and they are highly non-linear and non-stationary in nature. The pre-processing of the available epidemic data involves a thorough investigation of these non-linear and non-stationary features and an effort to eliminate them for the shake of simplifying the subsequent analysis.

The first 40 days confirmed daily and cumulative cases and deaths of COVID19 in each of several European countries including Greece, Germany and Switzerland, were extracted from the available databases (WHO, John Hopkins, ECDC). The data was cross-referenced to make sure that it is as accurate as possible. By comparing the deaths with the confirmed cases a first estimation of the Case Fatality Rate of each country (CFR) was derived. For the rest of this pre-processing step, a 5-day moving average was applied to the daily cases timeseries, in accordance with the statistical services standards.

The next goal was to be able to extract characteristic parameters of the disease from the available data. By using the active cases timeseries to extract a daily rate of epidemic increase, proportional to the growth rate, and fitting several functions from the family of gamma distributions, we managed to estimate the mean and the 95% CI recovery and infectious periods of the SARS-COV2 infection. More precisely, 600 different combinations of the scale and shape parameters of the Erlang distribution were regressed to the data of growth rate and the optimal distributions were selected using Akaike’s information criteria and Mean Squared error minimization. The characteristic recovery period is estimated to be 13.4 days (10.6, 18.9 95% CI), while the infectious period is estimated to be 8.4 days(5.5, 11 95% CI). These results are not terminal, in the sense that they differ from country to country. The safest possible estimation is provided in the results section.

### 3.3. Estimating effective reproduction number, daily transmissivity and CFR

One of the basic parameters of any epidemic is the basic reproduction number, known as Ro. Alongside, the effective reproduction number, which is the temporal variation of the reproduction number, they both describe the potential evolution of an epidemic spread in a community. In the short range, daily increments are used. The estimation of the effective reproduction number is performed backwards, using the probability density function of the optimal gamma distributions fitted to the data and the smoothed timeseries of the new cases. In order to capture variations in the progress of the infections among a population, the shape of the distribution as well as the duration of the infectious period were varied within the optimal range.

The basic equation used to derive Rt is:

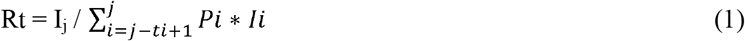

Where I_j_ is the n-day moving average of new cases and Pi is the normalized gamma probability density function at time i. t_i_ is the maximum infectious period, set at 10 days. The moving average is calculated for a 10-day time window (i.e. n=t_i_=10).

Both the daily cases and the constructed Rt timeseries were subjected to several non-stationarity tests. For this purpose, both the autocorrelation and partial autocorrelation functions were applied to the timeseries. The results showed that the new cases timeseries is probably non-stationary even in the short range, while the Rt timeseries is stationary in a wide sense. Therefore, several classes of ARMA models were fitted to the reconstructed 30-day Rt timeseries, in order to extend the timeseries to the forecasting period. 10 possible scenarios of the evolution of Rt with time were derived.

The transmissivity is the rate at which new infections are born from a susceptible population. It is directly proportional to the Ro. More details about the derivation of transmission rate are provided in the next subsection.

For a temporal extension of the CFR, the same method as that for the derivation of Rt is applied. Several ARMA models are fitted to the timeseries, as well as a GARCH model to evolve the error variance and add a bias to the timeseries.

### 3.4. Compartmental modelling

A double stage extended version of the demographic SEIR model with Erlang distributed incubation and recovery periods has been used for the simulation of the epidemic spread to a specific population. The population under investigation is divided into five subpopulations, i.e. Susceptible, Exposed, Infectious, Quarantined and Recovered. The infectious compartment is being subdivided into two different compartments, i.e. the infectious individuals at the onset of symptoms and the quarantined infectious individuals. In this model version quarantine involves both the fully isolated and the hospitalized patients, so that the model does not derive the two subsets separately and considers them as a single compartment. The fatalities are subtracted from the quarantine compartment using the timeseries of CFR as estimated by the aforementioned methods. An individual cannot simultaneously exist in two different subpopulations. The five subpopulations add up to the total population of the country under study. We also take into account the demographic features of each state, i.e. the birth and death rates.

Let t be the instant moment representing a specific day. The time increment is also daily. Let τ_σ_ denote the mean incubation period and τ_γ_ denote the mean recovery period. Let β represent the transmissition ratio, which is proportional to the basic reproductive number and vice versa. In general, the mean transmission ratio is linked to the basic reproductive number by the relationship:

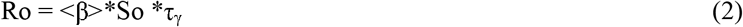

Where So is the initial susceptible population.

Alternatively, this relationship can be written as:

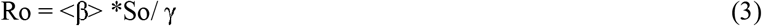

Where γ = 1/ τ_γ_ is the recovery ratio.

In an analogous manner, we may determine the incremental transmission ratio as:

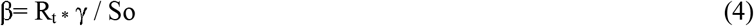

where Ro = < R_t_ >.

We also define the incubation ratio as σ=1/τ_σ_.

Now we are able to form the SEIR model equations, scaled in the (0,1) interval:

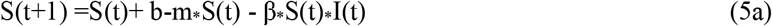

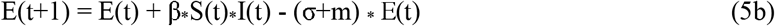

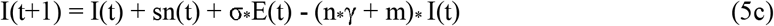

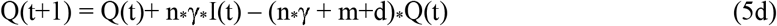

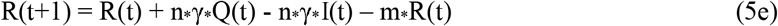

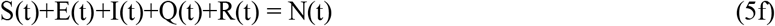

Where b : birth rate, m: natural death rate, d: CFR, n: scale of the Erlang probability distribution, sn(t): stochastic noise and N(t): the total population. For a compartmental SEIR model with two infectious compartments, n =2.

The original subpopulations are derived by scaling up the results to the total population, which is denoted as N_o_.

The equations are solved iteratively for the whole period of study, using the Euler’s method.

Last but not least, we need to define the initial conditions. For this purpose, we assume that a very small fraction of the population is initially exposed to the virus, while there are no infectious or quarantined individuals. Thus, the initial population compartments are:

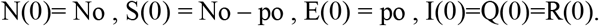

### 3.5. Sensitivity analysis

An intensive parametric study was contacted in order to derive the model parameters that seem to better describe the sequence of the recorded daily confirmed cases for the first 30 days since the first confirmed case. In order to validate the model, the estimated daily cases and deaths are compared with the recorded ones in terms of the mean absolute error (MAE) and the mean squared error (RMSE) normalized with the maximum daily cases recorded.

It was found that the characteristic times of the infection are dependent on the country under study and seem to be consistent with other similar studies. The median of the incubation period was estimated to be 5.1 days ([4.1, 6.1] 95% CI) and the median of the recovery period was estimated to be 17 days ([16, 18.5] 95% CI) for one version of the model and 14 days([12.5, 15] 95% CI) for another version of the model.

The basic reproductive number, Ro, which is the average of the daily reproductive number timeseries, was found to be unique for each country under study. For Greece, the median value at the first 3 weeks of the outbreak was estimated to be 2.92 ([2.72, 3.15] 95% CI). For the time period from the start of the infection to the lockdown withdrawal it was estimated to be 1.67 ([1.59, 1.76] 95% CI).

### 3.6. Model adjustment to the primary scientific goal

The model has been developed in several stages, each stage utilized for a different purpose. The pre-processing stage, which is the same for all versions, serves to estimate characteristic parameters of the disease, which are also used as parameters of the compartmental model. The compartmental model is utilized in two different ways. A deterministic and a stochastic. The deterministic version is used only for the short-term prediction of the epidemic spread. The model is initialized with specific initial conditions (the assumption used is that 100 people are initially exposed and 25 people are initially infectious to the virus) and “run” with a specific noise series, (check Appendix for details). A specific ratio of tested to real cases is assumed. The assumption made is that the quarantined individuals are 5 times more than the confirmed daily cases at all times, answering to a ratio equal to 0.2.

The stochastic model is used both for the prediction and the present estimation of the situation in terms of total cases, infectious, exposed, dead and recovered patients. The notion behind the stochastic model is to apply numerous different versions and try to reach to a specific conclusion. For this purpose, a novel Monte Carlo method is applied. 10 different scenarios of the daily reproduction number are combined with 1000 different scenarios about the time evolution of stochastic noise and 1000 different scenarios about the initially exposed individuals.

#### 3.6.1. Versions of stochastic noise

I. In the first version, the stochastic noise is assumed to be white noise with mean 1 and standard deviation equal to the standard deviation of the daily cases. The above versions offer 1000 random combinations of white noise and initially exposed individuals for each of Rt scenario.
II. In the second version, the stochastic noise is assumed to be a combination of two cosine functions, the frequency of each of which is derived from wavelet analysis of the sequence of new cases. In this case, the Monte Carlo scenarios are produced by varying the magnitude of the cosines according to the standard deviation of the noise contained in the signals. The basic form of the stochastic noise is derived from the dominant harmonic of the wavelet spectrum of new cases and the dominant harmonic of the wavelet spectrum of the noise extracted from the new cases:

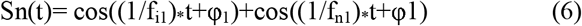

Where t is the time in days, f_i1_ =7 days, f_n1_ = 10 days and φ1=3 days.

The assumed ratio of tested to real cases is also allowed to vary within the interval [0.01, 0.5]. It varies 1000 times at a constant increment. This ratio has been selected not to affect the compartmental model, as the confirmed cases are derived from the model output at a secondary stage. The different versions of the white noise are produced by randomly generating numbers from the aforementioned normal distribution. The different versions of the initial conditions are produced by randomly generating positive integers at the [1,200] interval. The assumption made in this stage is that the maximum possible number of initially exposed individuals in Greece were 200 individuals and this value has been assumed by cross-referencing the official reports from EODY at the onset of the epidemic in Greece.

Therefore, the stochastic Monte Carlo compartmental model generates 10,000 scenarios about the evolution of the epidemic. The optimal Rt as well as the optimal combination of white / wavelet based noise and initial conditions are selected through a multi-stage MSE minimization process. First, the model selects 400 scenarios by MSE of deaths minimization. Those 4000 scenarios correspond to 4 out of 10 versions of Rt that best describe the available data and the 1000 combined white noise and initially exposed individuals scenarios for each of the 4 Rt timeseries. The optimal combination of Rt timeseries, white noise and initial conditions is then selected by minimizing a cost function, which is the sum of the MSE in terms of daily cases and the MSE in terms of deaths. By this process, the ‘best’ Rt timeseries as well as the best set of white noise and initial conditions versions can be selected off these 400 scenarios. An additional condition issued to the optimization is that the absolute error of total confirmed cases at the last timestep evaluated does not exceed 5%. The optimization leads to 4 × 25 reliable scenarios about the outbreak, from onset to lockdown withdrawal, while the ‘central scenario’ is the one that corresponds to the optimum Rt timeseries and the optimum combination of white noise and initial number of exposed individuals that corresponds to this Rt value. The mean of the 25 scenarios answering to the best Rt, is considered to be the most reliable index of the epidemic spread in the country. Alongside the minimum and maximum of these scenarios, it may provide a reliable range of the epidemic in terms of total infections, currently exposed, infectious and quarantined, expected total fatalities and recovered patients. Except fatalities, the estimation of the other variables primarily involves the true cases, while the confirmed cases are derived as a secondary task.

The stochastic model can also serve as an autonomous forecasting tool. By storing the results of the optimization, 30-day projections of the epidemic populations are produced. The only variable, the estimated values of which require alterations, is the reproduction number. We shall discuss about these changes in the next section. Referring to the information of the previous paragraph, it is obvious that 4 × 25 future scenarios about the course of the epidemic can be generated by running the compartmental model with the optimized parameters. In this way, an ensemble of forecasts is produced, the mean, maximum and minimum of which, may provide a reliable forecast. The central scenario is also included in the forecasts, while not being as reliable as the mean of the ensemble predictions.

## 4. Results

The estimated daily cases are derived from the infectious compartment. The quarantined compartment is used to make an estimation of the confirmed cases. For this purpose we assume that only a fraction of the quarantined individuals is tested. According to the Greek policy, only people with severe symptoms or links to abroad travelers are tested, so for the deterministic model the assumption made is that the confirmed cases are the 20% of the quarantined.

By using either the infectious or the quarantined compartment, we are able to construct an epidemic curve and also determine the temporal peak of the epidemic in each country. You should take into consideration that there is a lag between becoming infectious and quarantined owning to a time interval involving the incubation period and a small delay in the onset of symptoms. This phase difference is observed to be around 5-6 days. In addition, there is a lag between the epidemic curve constructed by quarantined individuals and confirmed daily cases, owing to the fact that there must be a time delay between onset of symptoms and confirming the cases by testing.

The total number of cases is derived by applying a cumulative function to the quarantined compartment. The observed difference between the cumulative infectious individuals and the cumulative quarantined individuals can be thought of as the asymptomatic proportion. According to study [11], the asymptomatic proportion is estimated between 17% and 33% of the total cases. This model seems to underestimate this proportion, but this is mainly due to the fact that we did not attempt to separately estimate the asymptomatic compartment.

### 4.1. Outcome of the pre-processing stage

It has been previously stated that the pre-processing stage is common for the deterministic and the stochastic model up to the stage where a stochastic noise is randomly generated. All these pre-processing stages involve the estimation of the mean infectious and recovery periods as well as of the daily variation of the reproduction number and of the Ro. For the derivation of these parameters, the first 40 days of the confirmed cases and deaths have been taken into account.

In order to derive the recovery time, the pre-processing algorithm was applied to 4 different European countries, i.e. France, Greece, Switzerland and Germany. The algorithm optimizes the shape and scale factors of the fitted gamma distributions. The mean recovery period is the inverse of the scale of the optimal gamma distribution. The algorithm produced different results for each country. For instance, in Switzerland, the mean recovery period was found to be 13.4 days (95% CI days: 10.6, 18.2), while in Germany it was found to be 15.4 days(95% CI days : 11.6, 20) and in Greece 19.7 days (95% CI days: 13.4, 20). It is believed that this inconsistency is partly a real situation due to the difference in race, health system and age of the patients and partly a data collecting issue. Due to reporting the data on time, the recovery period of Switzerland has been used as an input parameter for the rest of the pre-processing stage.

The algorithm is also capable of estimating the optimal shape factor of the gamma distribution, which is found to be equal to 4 for all countries. As for the infectious period, by definition, it is equivalent to the recovery period(the patient can infect from onset to symptoms till full recovery). However, as indicated by the gamma distribution, the probability of infecting others changes rapidly with time. It also needs to be considered that a patient may initially be infectious during a small period of time between the incubation and the onset of symptoms. This pre-symptomatic stage is estimated to be around 2 days. Last but not least, soon after the patient develops symptoms, he/she is either self-quarantined or hospitalized according to the country’s social distancing policy, thus not being able to infect others for the rest of the recovery period. Given these information, the infectious period is estimated to be around 5-6 days. However, for the estimation of Rt, the assumption that the infectious period varies between the confidence bounds of Switzerland is made. The representation of the social distancing and it’s effect on infectious stage duration is achieved by varying the shape factor of the gamma distribution.

More precisely, the optimal Rt is derived by assuming that the infectious period is 10 days at the most. The shape of the infection probability distribution is set at 2. In order to derive a range of possible Rt scenarios, the scale of the infection probability distribution is varied 10 times, using the range [10,19]. Notice the difference in the probability density functions of the infectious period for the different versions of the shape factor in the following graph:

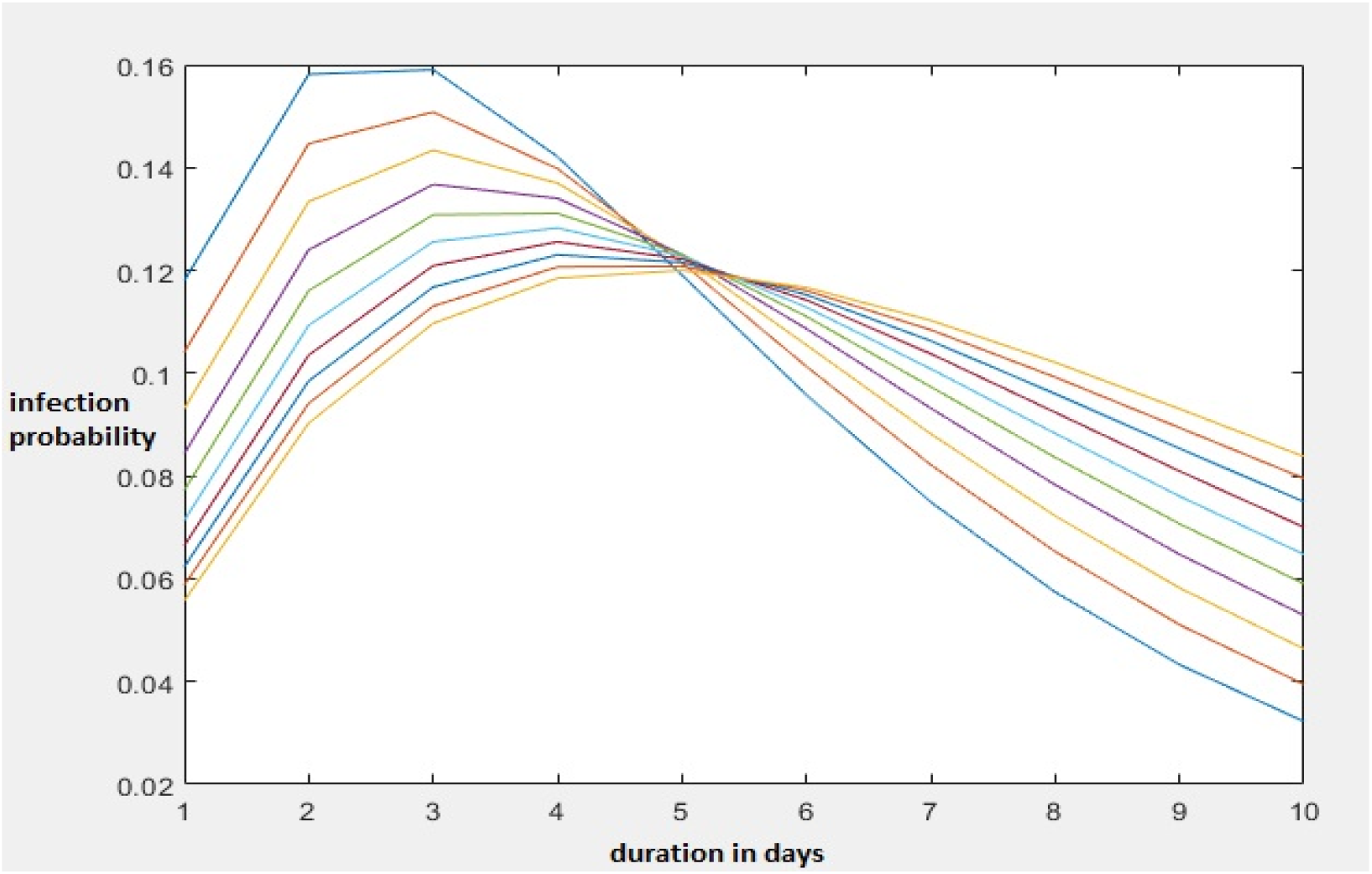

It is evident that as the scale factor increases, the infectious period becomes longer, but the maximum probability becomes shorter. The characteristic time varies from 6 to 18 days, but the upper limit of infectious periods is set at 10 days due to healthcare interventions.

For the estimation of the optimal Rt timeseries, a gamma distribution of shape factor of 2 and a unique scale factor equal to the inverse of the optimal recovery period(13.4 days) is used to estimate infection probabilities.

For the derivation of the various Rt scenarios used in the rest of the study, as aforementioned, distributions with the same shapes but varying scales are fitted to the data.

The optimal Rt timeseries for the 70 days of this study, from epidemic start to lockdown withdrawal is shown in the following graph:

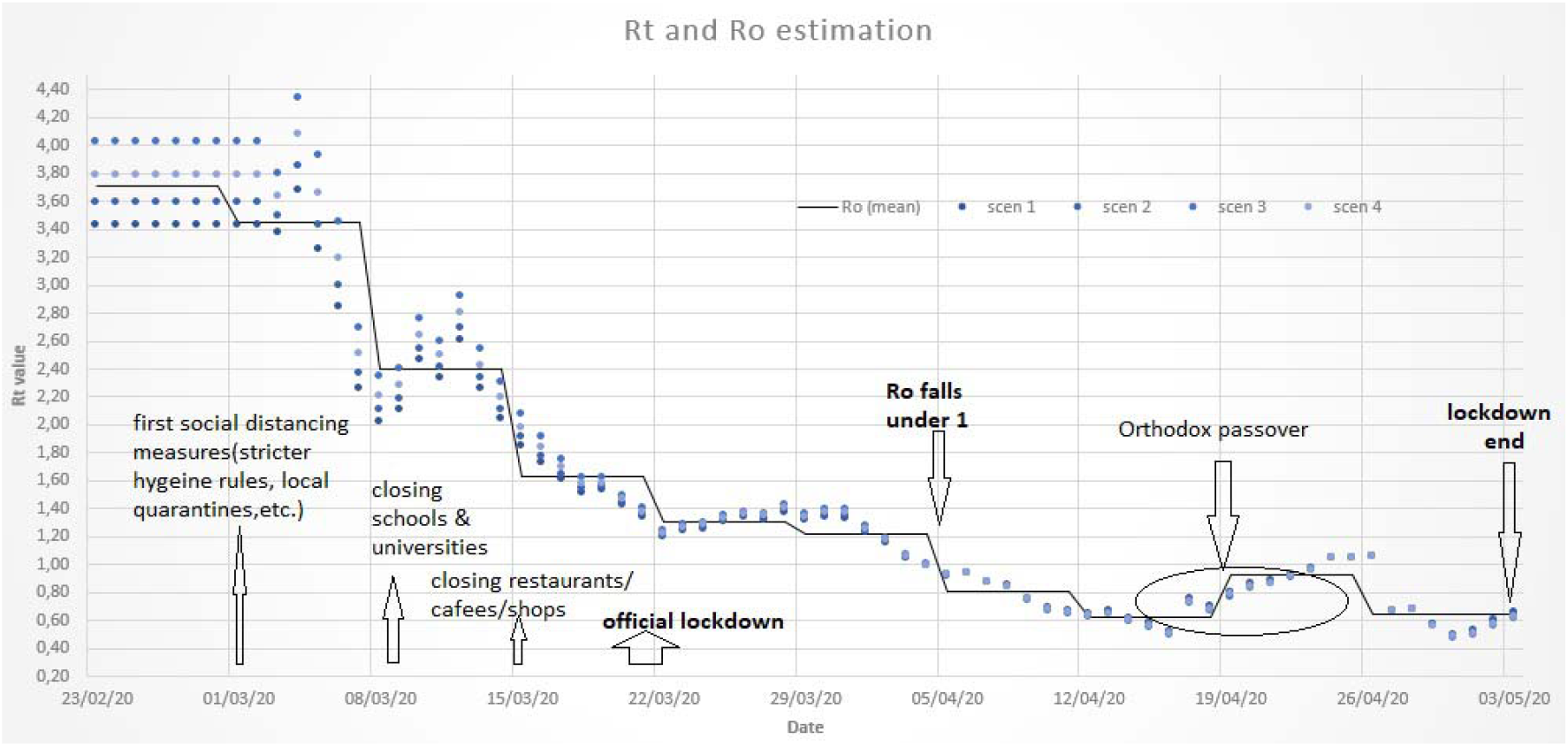

The Ro is inferred by averaging of the Rt sequence over 7 days. So, in this study, Ro is the weekly average of Rt. The estimated Ro values for each week of the 70-day period in study, is shown in the following table:

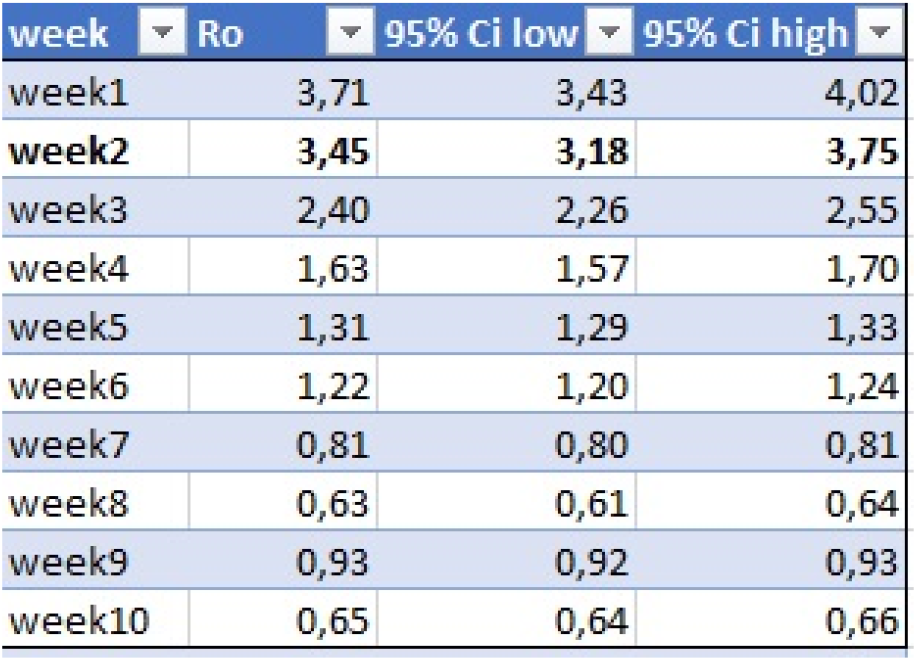

It is evident that Ro without interventions, started from a value around 3.7 but dropped quickly to a value around 3 during the first 20 days of the outbreak, while after the gradual application of the strict social distancing measures, it started dropping under 2 and 15 days later it started dropping under 1, indicating the epidemic decline. It can be safely assumed that the basic Ro value in Greece was 3.4 ([3.2, 3.8] 95% CI). A week later than the application of the first social distancing measures it dropped to a value of 2.4 (2.26, 2.55 95% CI). Closing schools and universities resulted in a further decline of the Ro to 1.6 at week 4 after the epidemic start, while closing shops, restaurants and cafees led to a further small reduction of the reproductive number at weeks 5 and 6. It is however evident that only after the official lockdown was decided, at the start of the 5^th^ week, the Ro started dropping under the critical value of 1. This was the start of the epidemic decline in Greece. The epidemic made an unsuccessful effort to re-attack during the Passover, a period during which several individuals tried to break the government measures, but the Ro never surpassed 1 and then it dropped again to a value very close to 0.5. At that time, the Greek government seems to have made the decision to gradually withdraw the lockdown.

### 4.2. Forecast validation of the deterministic model

The mean of the 10 Rt scenarios is used as the input for the transmissivity, while the recovery period is set at 17 days (the median value of the Greek confidence interval) and the incubation period is set at 5.1 days. Demographic information are included, while the noise timeseries is reconstructed by time-extending the inverse of the Fourier power spectrum of daily confirmed cases. The model is initialized with 125 exposed and 25 infectious individuals at day 1. The results of the compartmental model in terms of total confirmed cases and deaths are shown below:

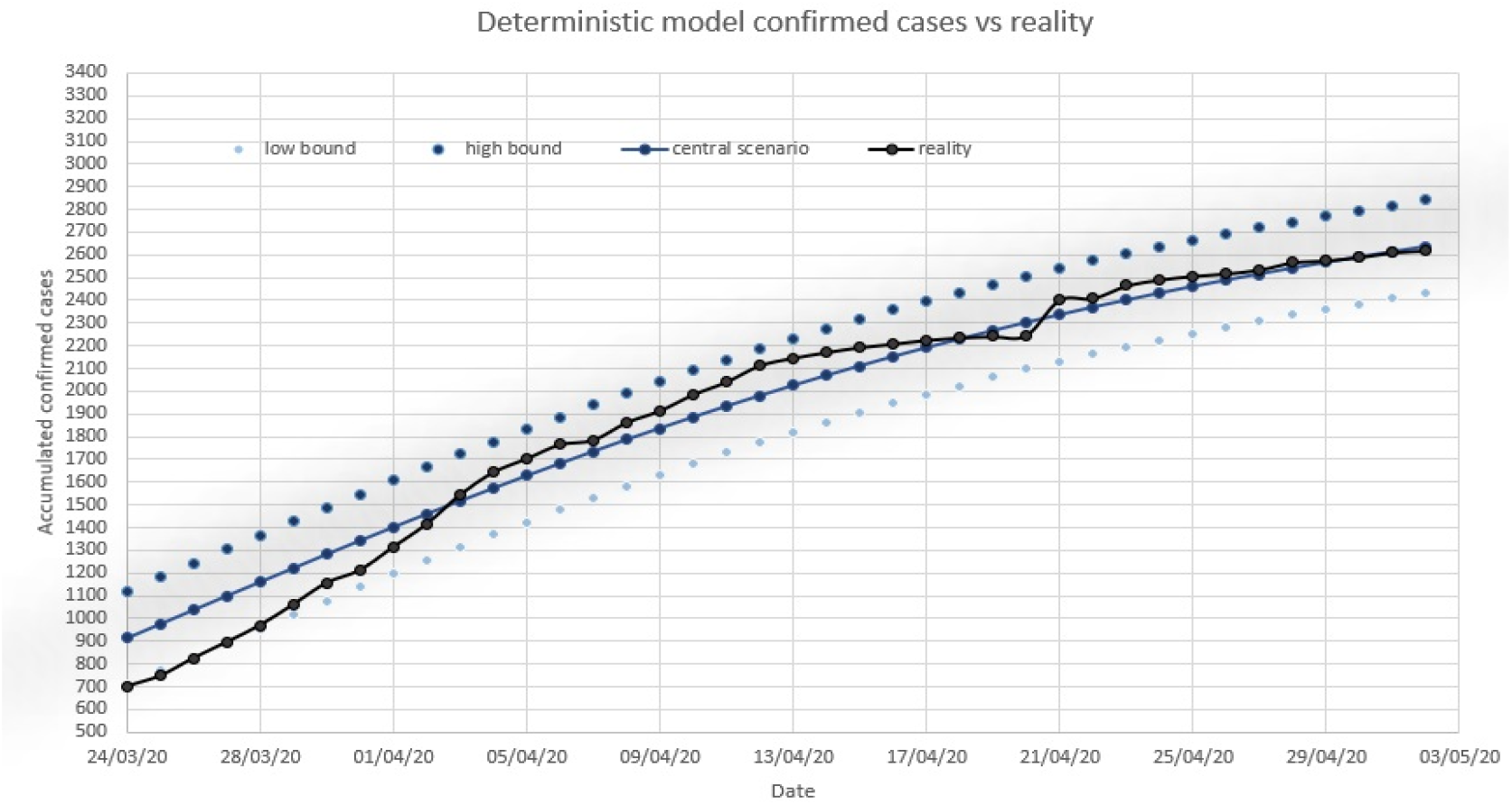

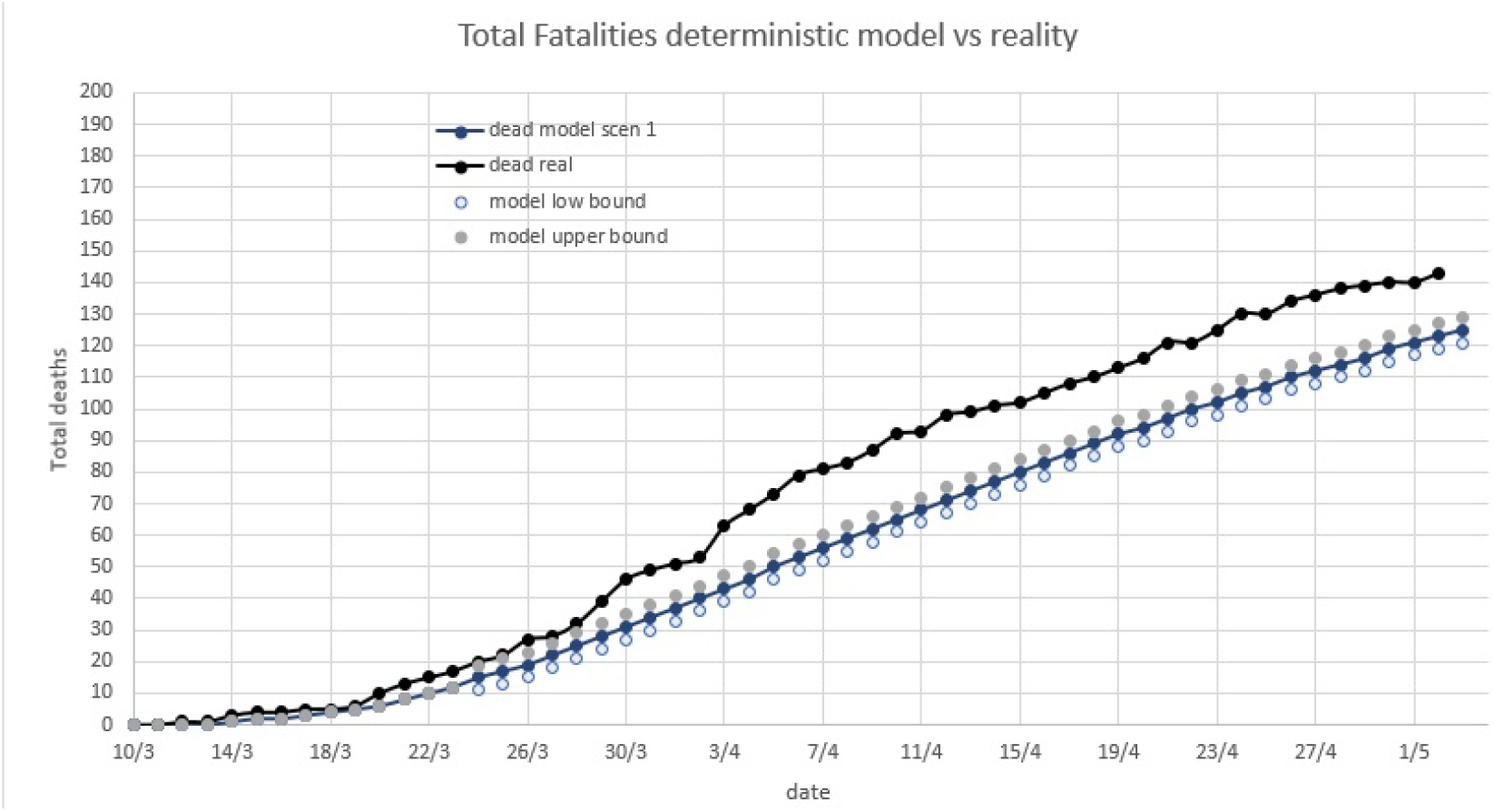

Although the model is initialised since the epidemic start, it was decided to validate the forecasts after the 35^th^ day, taking into account that real data of 40 days has been used in the pre-processing stage. Therefore, the validation of the forecasts has been performed for 35 days, i.e. from March 28^th^ to May 2^nd^. This decision applies to both the deterministic and the stochastic model. The MAE and RMSE error metrics, normalised by the accumulated number of cases at each timestep, are used to evaluate the forecasts.

In terms of confirmed cases, the MAE of the central scenario is 2.6%, while the RMSE is 3.1%. The 95% confidence bounds provide an accuracy of roughly +-8% with respect to the RMSE. The model has mostly captured the accumulation of confirmed cases, although there is a slight underestimation around the peak time of the epidemic and a systematic overestimation of cases near the lowest timesteps of the forecast.

In terms of confirmed deaths, the MAE of the central scenario is 13%, while the RMSE is 13.5%. The 95% confidence bounds provide an accuracy of roughly −15% with respect to the RMSE. The model systematically underestimated the fatalities.

### 4.3. Forecast validation and outcome of the stochastic model

#### I. White noise stochastic model

The ensemble forecast of the white noise stochastic model is validated for the same period as for the deterministic model. The results of the forecasts in terms of total confirmed cases, deaths and daily confirmed cases are shown below.

The central scenario, the ensemble mean, maximum and minimum are evaluated with respect to the confirmed cases/deaths.

In terms of total confirmed cases, the MAE of the central scenario is 3.57 %, while the RMSE is 4.3%. The MAE of the ensemble mean is 3.9%, while the RMSE is 4.6%. The ensemble min and max produced a forecast error of 7.2 % and 4.3% in terms of MAE and 8.4%, 5.4% in terms of RMSE, respectively. It is evident that the stochastic model has better captured both the short-term and the long-term variations and magnitude of the cases than the deterministic model and the forecast is almost entirely within the ensemble range. The central scenario has performed slightly better than the ensemble mean.

In terms of total fatalities, the central scenario shares a MAE of 5.6% and an RMSE of 6.4%, while the ensemble mean’s MAE is 15.3 % and the ensemble mean’s RMSE is 15.8%. The superiority of the stochastic model with respect to the deterministic one is clearly evident for this variable, having achieved an accuracy improvement of more than 60%.

By comparing the real and forecasted epidemic curves, it is obvious that the model has managed to forecast the temporal peak of the epidemic in Greece by a deviation of less than 3 days. Recall however that the real epidemic curve is constructed from the confirmed daily cases, so there is a justified lag between the new quarantined patients estimated by the model and the confirmed tests. On the other hand, the model failed to capture the little details with respect to daily variations of the cases announced. This indicates that a wavelet analysis could improve this inaccuracy.

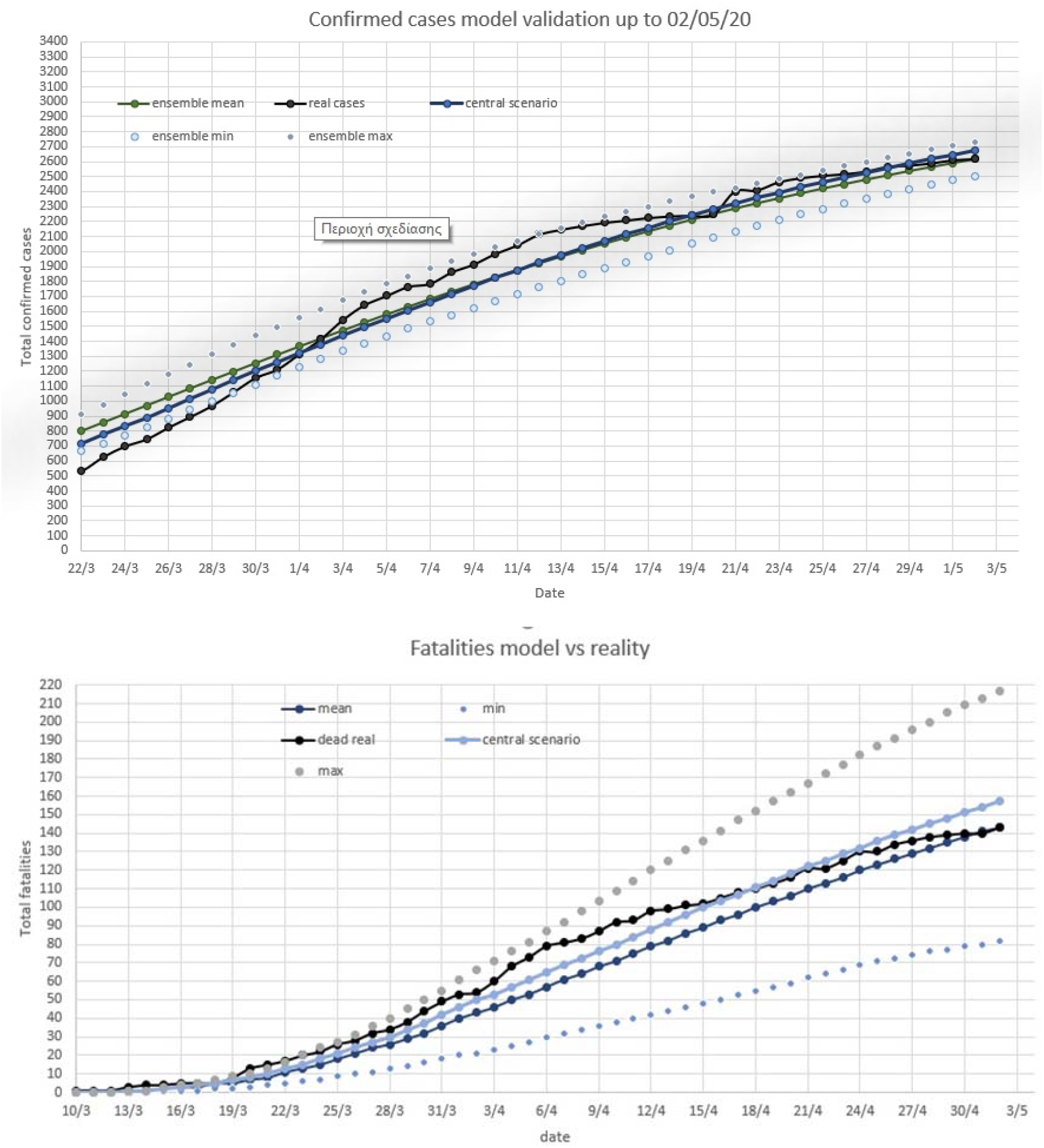

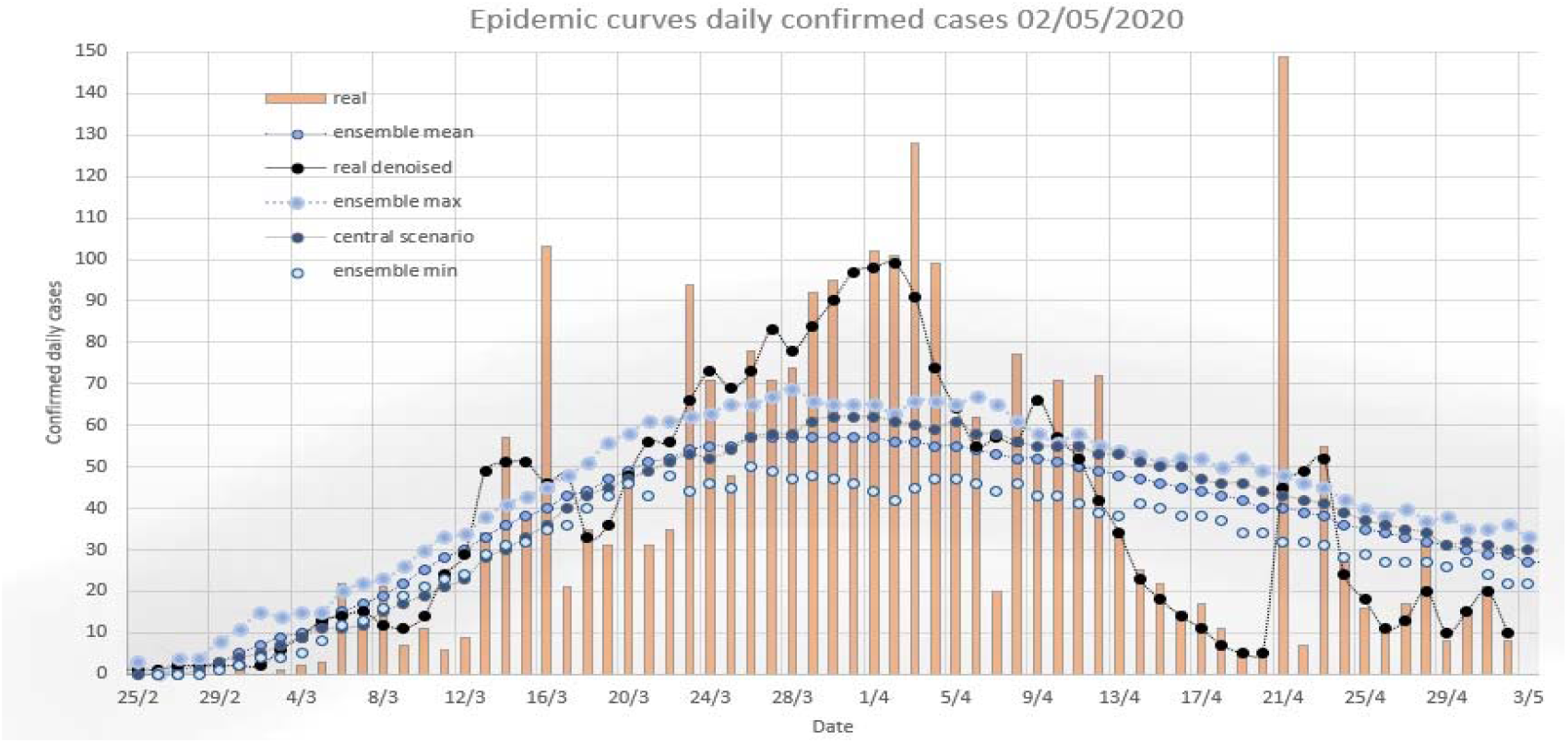

### Estimation of the real magnitude of the epidemic in Greece

The outcome of the ensemble system of compartmental models in terms of accumulated and daily cases, as extracted from the quarantined compartment, is shown in the following graphs.

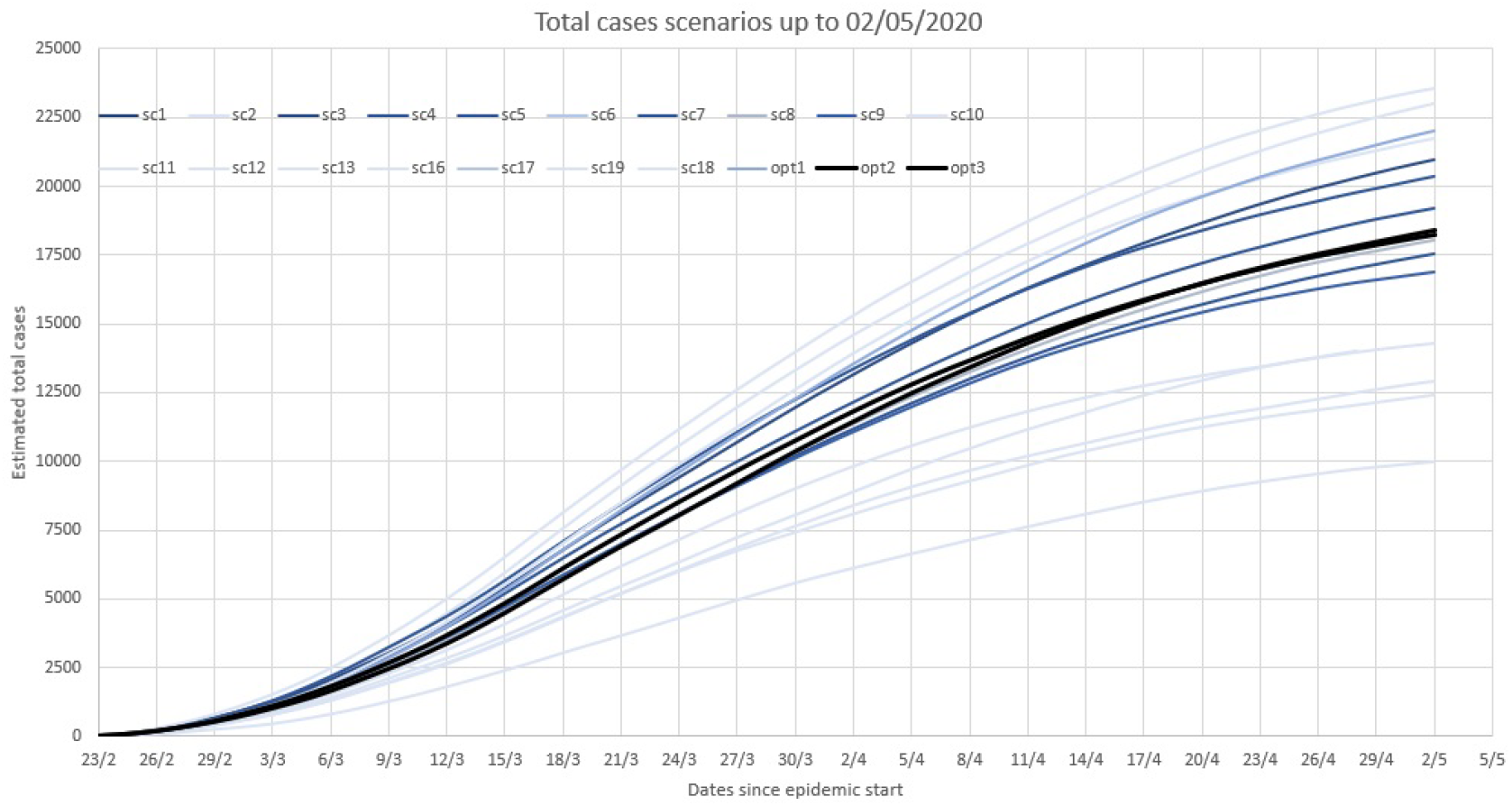

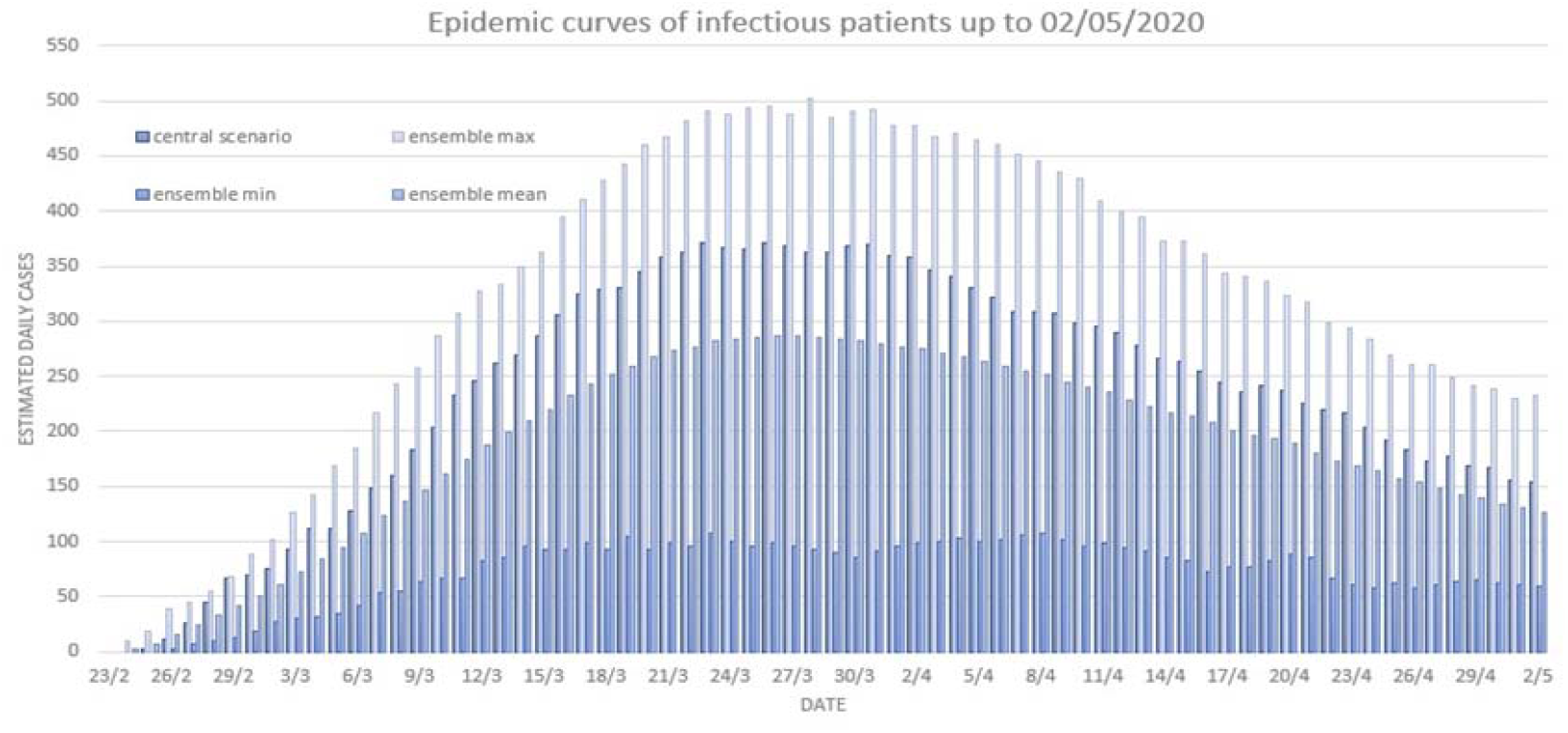

The period under study is from the onset of the epidemic in Greece to the last day of the lockdown. Total cases estimation includes all infectious individuals, irrespective of their severity of symptoms, including fatalities. The epidemic curve involves infectious individuals who have been quarantined and not died.

In order to estimate the real magnitude of the epidemic, the full ensemble scenarios for the 4 best guesses of the Rt timeseries are incorporated to the analysis carried out hereafter.

It is assumed that the outputs of these scenarios follow a normal distribution with mean equal to the average of the ensemble outputs and standard deviation equal to the standard deviation of the ensemble outputs.

By the time of lockdown ending :

The median value of the estimated total cases scenarios is 12700. The maximum of the estimations is 23500 and the 95^th^ percentile is 22000 cases, i.e. there is a 95% chance that the total cases were equal to or less than 22000. Also, there is a 10% chance that the total cases were less than 5200. 60% of scenarios lies between 11200 and 23500 cases. Taking the assumption of normal distribution of the scenarios for granted, it follows that the upper statistical bound of the total infected individuals is 29950 cases.

Taking the optimal scenario of each of the 4 versions of Rt timeseries, the most reliable range of total cases is estimated between 18000 and 19000.

By the time of the epidemic peak:

The median value of the estimated scenarios total is 8100. The maximum of the scenarios is equal to 15000 infected individuals and the 95^th^ percentile is equal to 14500 cases. There is a 10% chance that the total cases were less than 3100. 60% of the scenarios lies between 7100 and 15000 cases.

The optimal range of total infected individuals by the time of the epidemic peak is estimated to be 11000-12000.

#### II. Wavelet-based noise stochastic model

The ensemble forecast of the wavelet-based noise stochastic model is validated for the same period as for the deterministic and the white noise stochastic model. The results of the forecasts in terms of total confirmed cases, deaths and daily confirmed cases are shown below.

The central scenario, the ensemble mean, maximum and minimum are evaluated with respect to the confirmed cases/deaths.

In terms of total confirmed cases, the MAE of the central scenario is 3.27 %, while the RMSE is 3.94%. The MAE of the ensemble mean is 3.61%, while the RMSE is 4.25%. The ensemble min and max produced a forecast error of 5.8 % and 3.8% in terms of MAE and 6.3%, 4.6% in terms of RMSE, respectively. It is evident that the wavelet-based stochastic model has slightly better captured both the short-term and the long-term variations and magnitude of the cases than the white-noise stochastic model and the forecast is almost entirely within the ensemble range. The improvement achieved by the wavelet analysis is mostly evident from the smaller RMSE values. The central scenario has performed slightly better than the ensemble mean.

In terms of total fatalities, the central scenario shares a MAE of 6.4% and an RMSE of 7.1%, while the ensemble mean’s MAE is 17.3 % and the ensemble mean’s RMSE is 17.8%. The superiority of the wavelet-based stochastic model over the deterministic one is clearly evident for this variable, having achieved an accuracy improvement of more than 60%. However, the white noise model has performed slightly better than the wavelet model in terms of deaths.

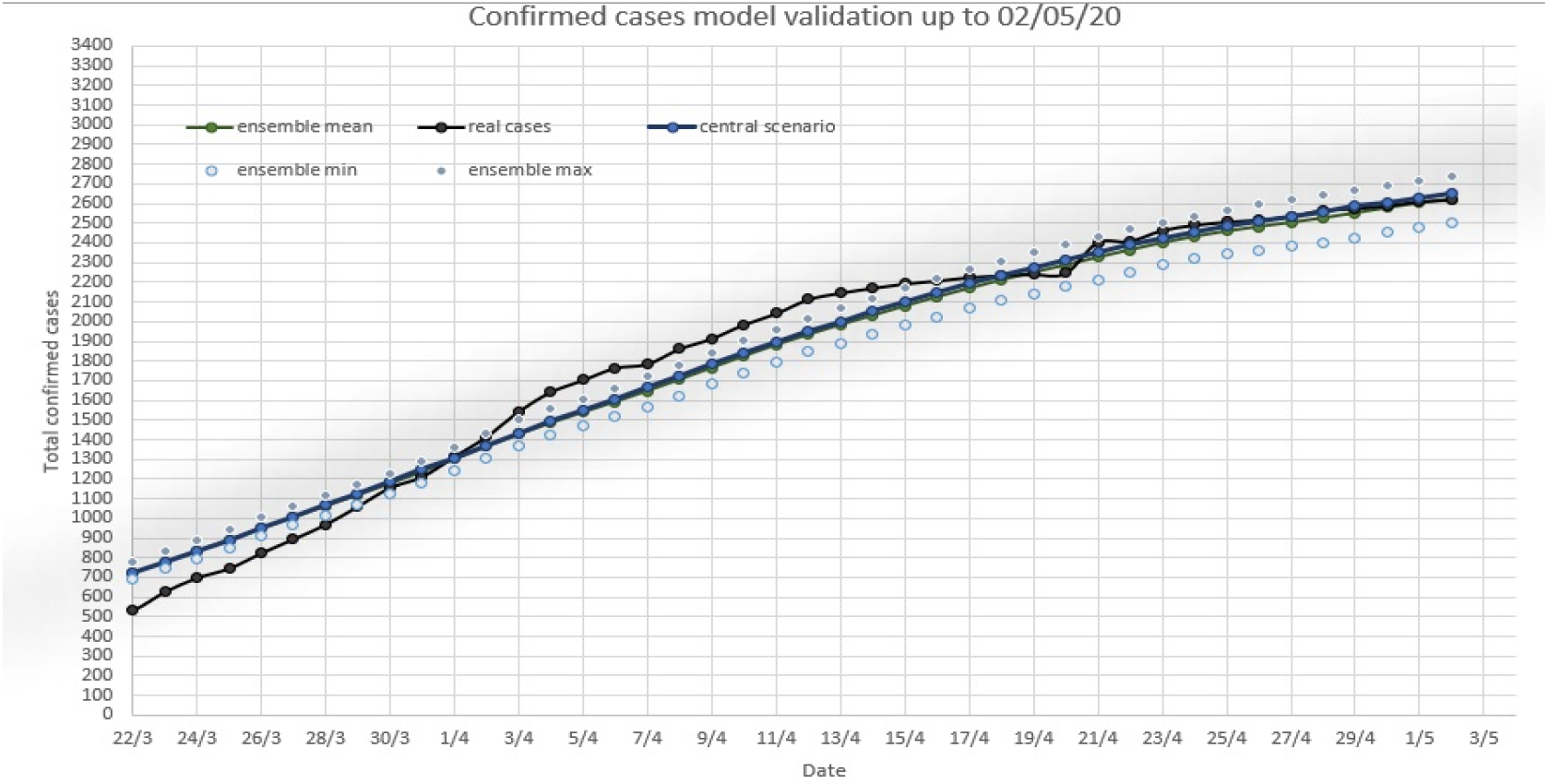

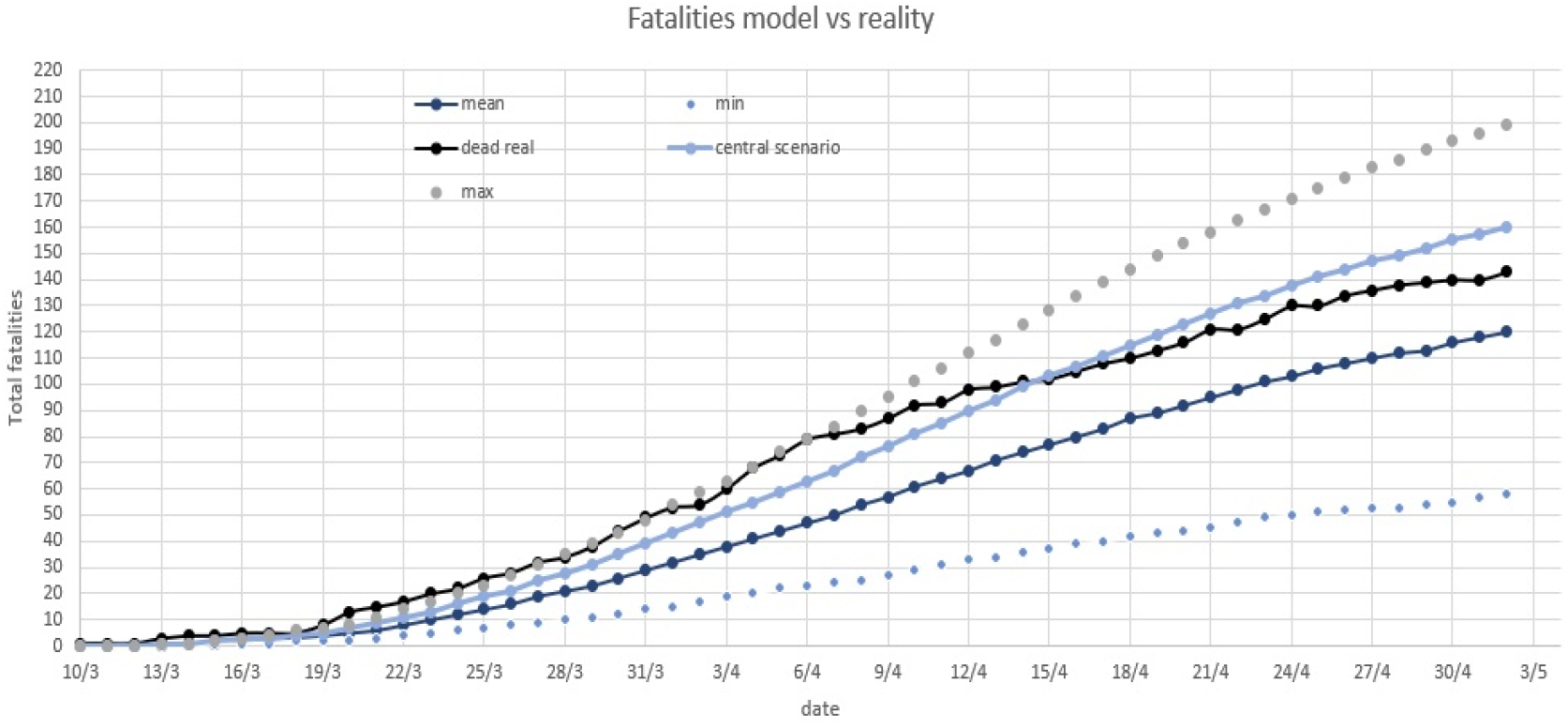

The most important improvement of the wavelet-based model over the white-noise model is in terms of the depiction of the epidemic curve of confirmed cases. As indicated by the graphs, the wavelet analysis is able to better capture the temporal variations of the epidemic as well as the magnitude of the spikes and the reconstructed epidemic curve clearly follows the real one.

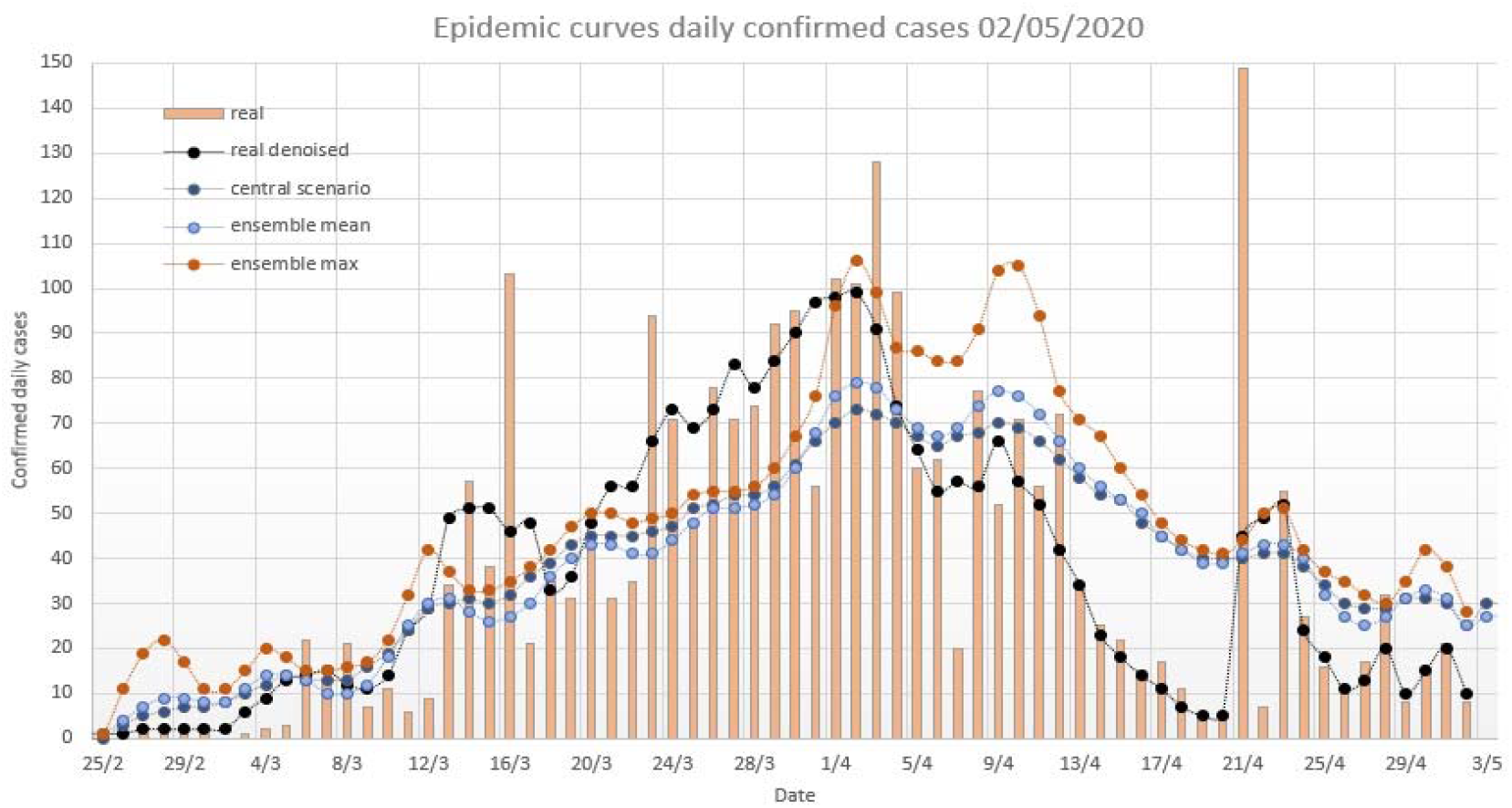

As for the real magnitude of the epidemic, similar results with the white noise model were obtained. By the time of lockdown ending:

The median value of the estimated total cases scenarios is 12000. The maximum of the estimations is 22900 and the 95th percentile is 20900 cases, i.e. there is a 95% chance that the total cases were equal to or less than 20900. Also, there is a 10% chance that the total cases were less than 8100. 60% of scenarios lies between 11000 and 22900 cases. Taking the assumption of normal distribution of the scenarios for granted, it follows that the upper statistical bound of the total infected individuals is 28950 cases. The optimal scenarios are estimated to be between 16000 and 17000 cases.

### 4.4. Future projections and performance of the ensemble system

For the purpose of forecasting the potential evolution of the epidemic after the lockdown withdrawal, the 4 best scenarios of Rt evolution as well as 25 ensemble scenarios for each of the 4 Rt timeseries are selected as initial conditions. The initial exposed, infectious and quarantined individuals of the projections are the last time interval output of the stochastic compartmental model. The noise is projected according to the best selected scenarios. This process produces 4x 25 30-day forecast ensemble scenarios, the mean, maximum and minimum values of which are included in the graphs. The central scenario is the one corresponding to the optimal combination of Rt timeseries and stochastic noise. For the projection of the Rt timeseries, it is assumed that the transmissivity follows a stepwise increase, where each step corresponds to the 1^st^, 2^nd^, 3^rd^ and 4^th^ week of the particular forecast. This assumption follows the official announcements on the social distancing measures withdrawal, which is also held step by step.

#### I. White noise stochastic model

The 30-day ensemble forecast in terms of new confirmed cases, cumulative confirmed cases and total fatalities along with the real values of these variables are shown below:

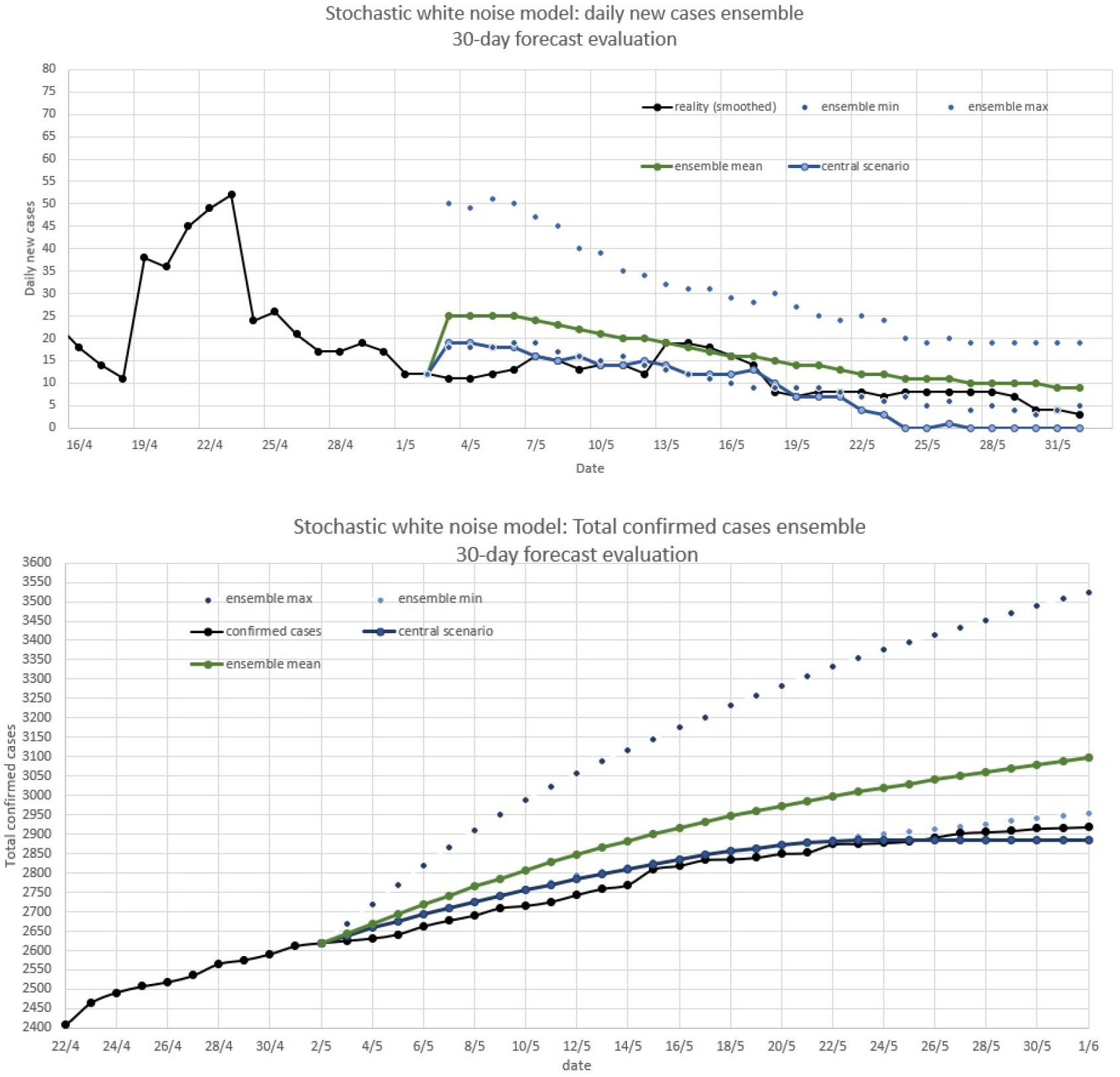

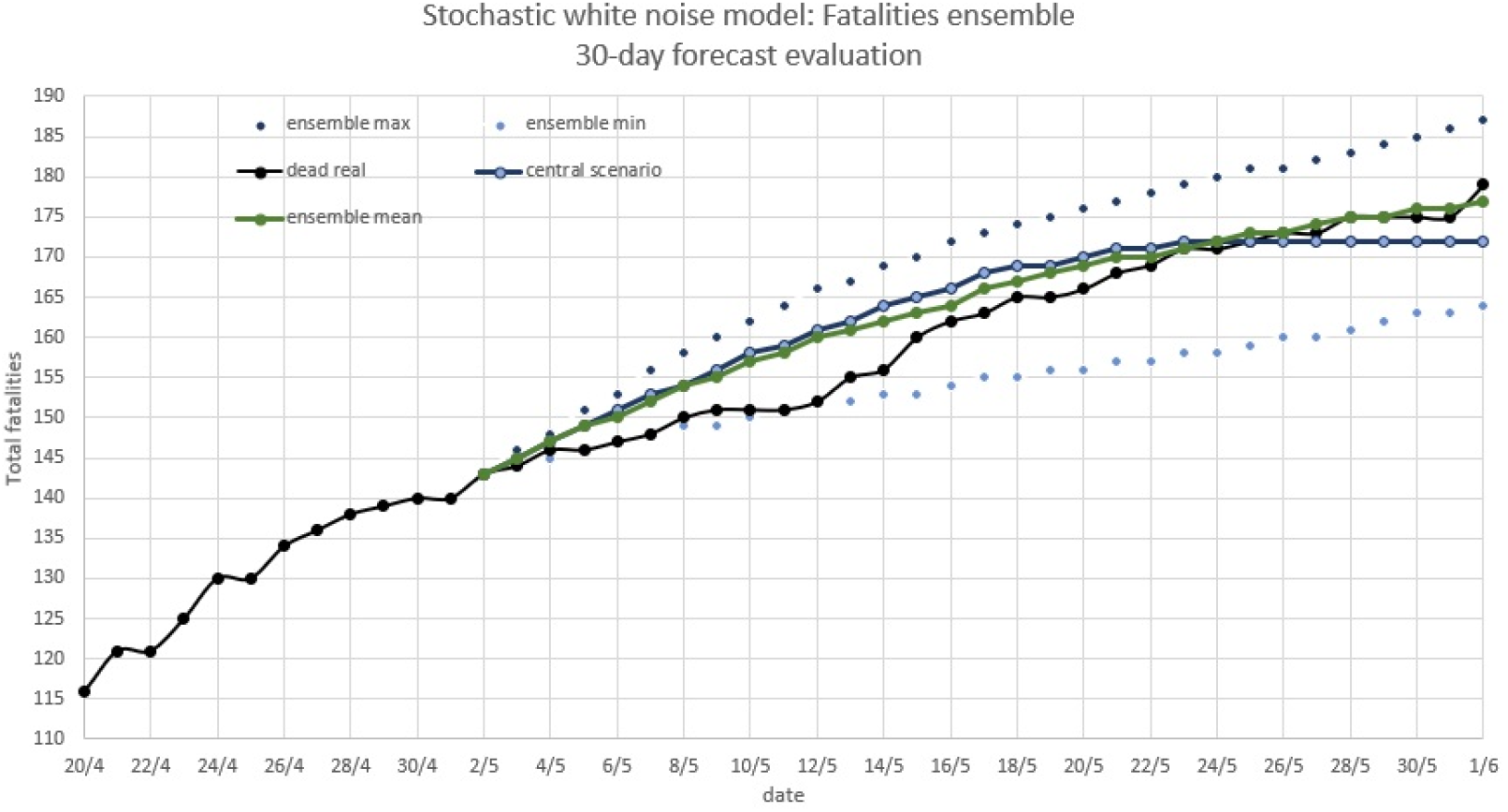

The ensemble system has very well captured the evolution of the epidemic during the last month. It cleary showed the gradual decline in new cases, but slightly overestimated the fatatilies during the first ten days. In terms of the total cases, the average of the ensemble scenarios scored a MAE of 3.9% and an RMSE of 4.2%, while the central scenario scored a MAE of 0.86% and an RMSE of 0.92%. In terms of total fatalities, the mean of the scenarios scored a MAE of 1.6% and an RMSE of 1.9%, while the central scored a MAE of 2.4% and an RMSE of 2.5%. With respect to new daily cases, it is evident from the corresponding graph that the white noise ensemble system has captured the general trend, but has failed to capture the short-term variability in the time series. Also, the central scenario has significantly underestimated the new cases, especially during the second half of the forecasting period, while the average of the scenarios has performed better.

#### II. Wavelet-based noise stochastic model

The 30-day ensemble forecast in terms of new confirmed cases, cumulative confirmed cases and total fatalities along with the real values of the variables, are shown below:

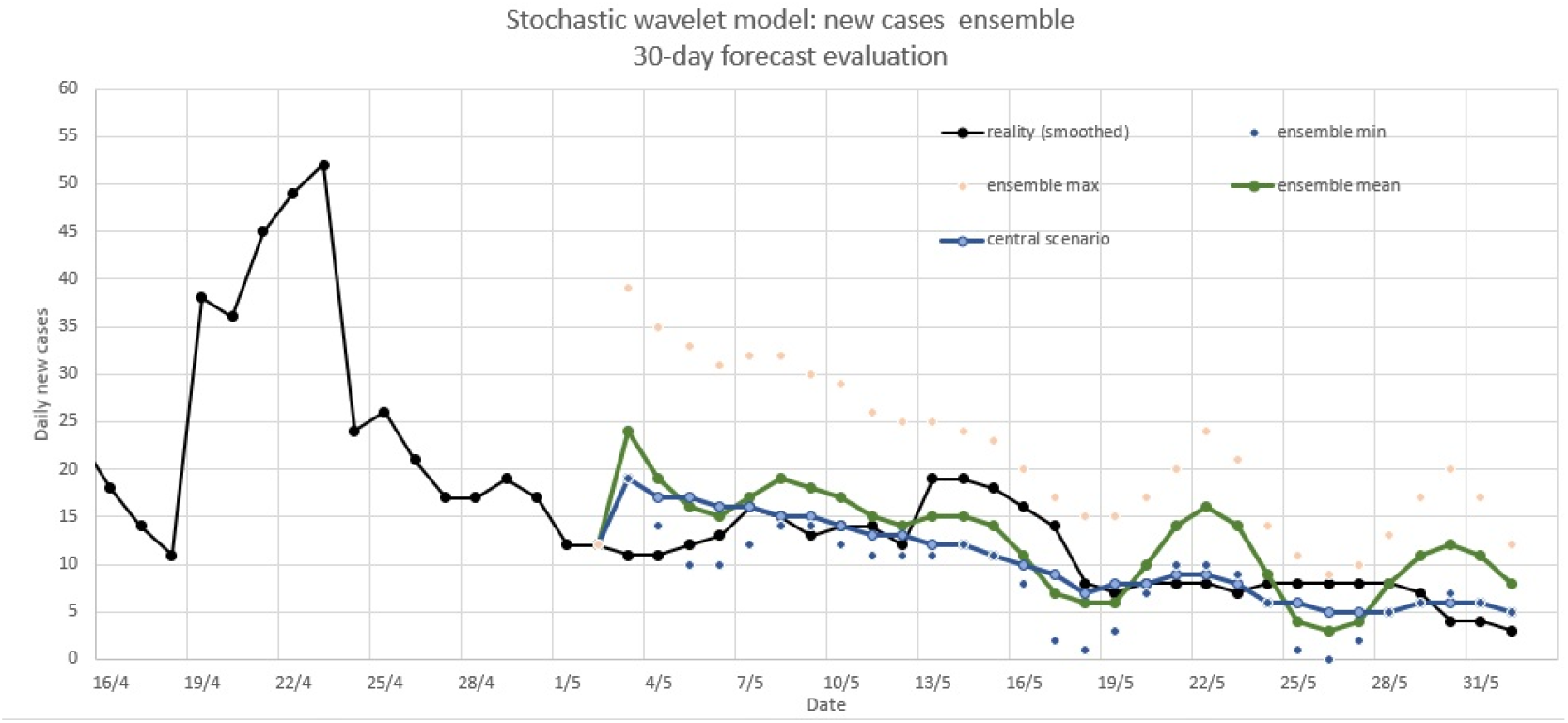

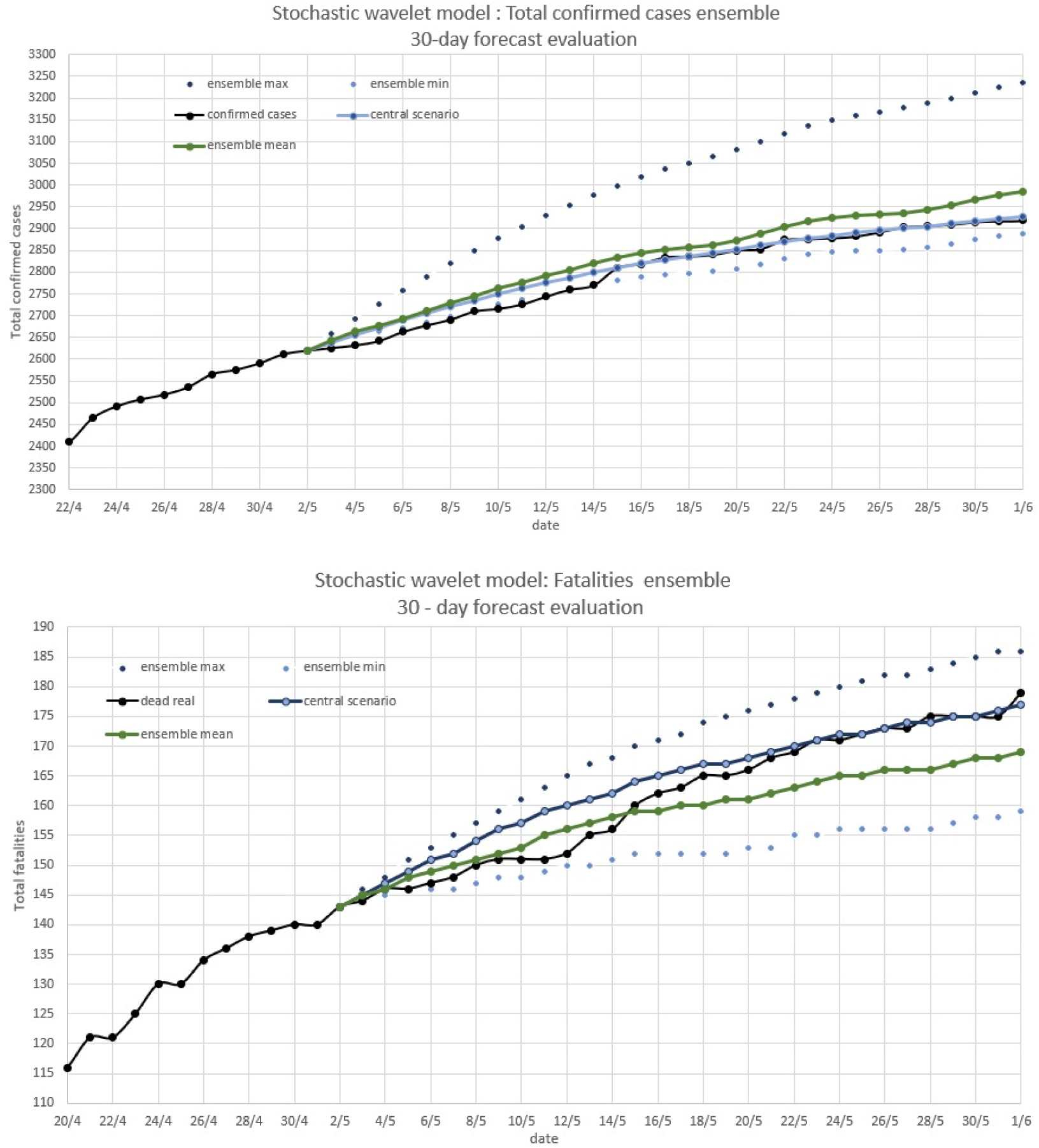

The wavelet based ensemble system has also very well captured the evolution of the epidemic in Greece after the lockdown withdrawal, showing clearly the decline of the outbreak. It also captured short-term variability in the confirmation of new cases. In terms of the total cases, the average of the ensemble scenarios scored a MAE of 1.26% and an RMSE of 1.28%, while the central scenario scored a MAE of 0.47% and an RMSE of 0.62%. In terms of total fatalities, the mean of the scenarios scored a MAE of 2.4% and an RMSE of 2.6%, while the central scored a MAE of 1.68% and an RMSE of 1.95%. With respect to new daily cases, the wavelet-based model captured both the trend and the short-term variability. Overall, the wavelet-based model outperformed the white-noise model by a margin of 20%. The superiority of the wavelet-based model is more profound in the prediction of new cases than in the prediction of new deaths.

## 5. Summary and Discussion

In this study, a variety of novel stochastic compartmental models of the Covid19 pandemic were developed for Greece. The mathematical basis of the models is a SEIR model with two infection stages. Two different noise schemes were incorporated to the model. The first is a white-noise with mean 1 and standard deviation equal to 6 and the second is wavelet-based noise, reconstructed as a sum of two cosines. In both schemes, the noise was varied randomly in order to create Monte Carlo simulations. The initial conditions were also varied randomly, while the ratio of tested to real cases was assumed to increase uniformly from 0.01 to 0.5 at each Monte Carlo scenario. By using a simple optimization algorithm, the model managed to safely estimate the real magnitude of the virus outbreak in Greece for the time-period between the start of the epidemic and the lockdown withdrawal as well as for the 1^st^ month after the lockdown withdrawal. It also managed to accurately forecast the evolution of the epidemic both in terms of confirmed cases and fatalities. Recovered individuals and active infections can also be inferred from the model output.

The wavelet-based cosine noise stochastic model outperformed the white-noise model in terms of the 30-day forecast of new and accumulated confirmed cases. In terms of total fatalities, the two models shared similar scores.

Through the estimation of the real magnitude of the epidemic in Greece, one can infer the ratio of tested to real cases, the actual fatality rate and the immunity rate. The ratio of tested to real cases is estimated to be between 0.12 and 0.18, which means that the total infected individuals are 6 to 9 times more than the confirmed ones. Subsequently, the actual fatality rate (IFR) is estimated to be 1.2% ([0.7, 2.1] 90% CI) and the immunity till June 1^st^ is around 0.17% of the total population ([0.12, 0.23]% 90% CI).

Both stochastic models can serve as valuable forecasting tools for healthcare services via the well validated ensemble system they provide. Robust and accurate predictions of up to 30 days and fairly accurate predictions of up to 60 days can be generated by the models. As more data is added to the model, more robust estimations of the characteristic parameters and the outbreak of Covid19 can be performed. Future work could focus on the inclusion of Markov chain probabilities as a substitute for the basic compartmental models used. Also, urban mobility patterns may be used to optimally infer the incremental reproduction number and the transmissivity. Modelling the effect of stochasticity is a subject of ongoing research, with more and more promising results, capable of boosting our understanding of infectious diseases.

## Data Availability

I hereby declare that all data and code scripts used for the conduct of this research are available if requested. I claim my mental rights, in case the ideas and methods used are copyrighted or used by another researcher.

